# AI in Medical Education: A Comparative Analysis of GPT-4 and GPT-3.5 on Turkish Medical Specialization Exam Performance

**DOI:** 10.1101/2023.07.12.23292564

**Authors:** Mustafa Eray Kılıç

## Abstract

**Background/aim:** Large-scale language models (LLMs), such as GPT-4 and GPT-3.5, have demonstrated remarkable potential in the rapidly developing field of artificial intelligence (AI) in education. The use of these models in medical education, especially their effectiveness in situations such as the Turkish Medical Specialty Examination (TUS), is yet understudied. This study evaluates how well GPT-4 and GPT-3.5 respond to TUS questions, providing important insight into the real-world uses and difficulties of AI in medical education.

**Materials and methods:** In the study, 1440 medical questions were examined using data from six Turkish Medical Specialties examinations. GPT-4 and GPT-3.5 AI models were utilized to provide answers, and IBM SPSS 26.0 software was used for data analysis. For advanced enquiries, correlation analysis and regression analysis were used.

**Results:** GPT-4 demonstrated a better overall success rate (70.56%) than GPT-3.5 (40.17%) and physicians (38.14%) in this study examining the competency of GPT-4 and GPT-3.5 in answering questions from the Turkish Medical Specialization Exam (TUS). Notably, GPT-4 delivered more accurate answers and made fewer errors than GPT-3.5, yet the two models skipped about the same number of questions. Compared to physicians, GPT-4 produced more accurate answers and a better overall score. In terms of the number of accurate responses, GPT-3.5 performed slightly better than physicians. Between GPT-4 and GPT-3.5, GPT-4 and the doctors, and GPT-3.5 and the doctors, the success rates varied dramatically. Performance ratios differed across domains, with doctors outperforming AI in tests involving anatomy, whereas AI models performed best in tests involving pharmacology.

**Conclusions:** In this study, GPT-4 and GPT-3.5 AI models showed superior performance in answering Turkish Medical Specialization Exam questions. Despite their abilities, these models demonstrated limitations in reasoning beyond given knowledge, particularly in anatomy. The study recommends adding AI support to medical education to enhance the critical interaction with these technologies.

## 1. Introduction

ChatGPT and other large-scale language models (LLMs) indicate a new era in natural language processing (NLP), a branch of artificial intelligence (AI) that allows human-computer interaction through natural language [1, 2]. The capacity of these models to recognize, analyze, and create human speech has the potential to alter numerous industries, including education, healthcare, customer service, and marketing [3]. The refining and evolution of these models have occurred over time, hastening the current technological revolution.

The use of LLM in education has attracted a great amount attention. LLM is used to improve teaching and learning processes, which has resulted in the creation of applications such as automatic essay scoring, plagiarism detection, intelligent tutoring systems, and language learning apps [4, 5, 6]. The significance of these apps in medical education cannot be overstated. Providing options for learning support, student grading, and curriculum updating to improve the quality of medical education while lowering expenses [7].

Although we are limited to using LLMs in medical education, complex models like OpenAI’s GPT-3.5 and its sequel GPT-4 have emerged as game changers. GPT-3.5, an improved version of the previous version, was used to build personalized learning materials, feedback on student achievement, and even complete courses [8]. GPT-3.5’s performance has been assessed and verified in peer-reviewed papers, indicating its capacity to create high-quality instructional content similar to human experts [9].

The launch of GPT-4, which is supposed to be more effective than its predecessor, caught the curiosity of specialists. GPT-4 has the ability to take medical education to unparalleled heights by comprehending and creating more complex languages and accomplishing more difficult tasks [10]. However, using these AI models in medical education poses obstacles. Models should be used ethically and responsibly, while respecting the privacy and autonomy of healthcare professionals and patients and avoiding bias and the propagation of prejudice [7, 11].

On the other hand, the idea of utilizing AI to help learning for medical students is projected to increase training quality at a low cost [12]. However, incorporating AI into traditional medical education is a challenging process fraught with difficulties such as determining AI efficacy and the technological complexities required in designing AI applications.

The process of evaluation and testing is an essential component of medical education. The Turkish Medical Specialty Examination (TUS), a key milestone in the professional growth of Turkish doctors with specific education, is one such evaluation in the Turkish medical school system. The TUS is a national test that assesses a specialist’s comprehensive knowledge and skills [13]. It is separated into two sections: clinical and basic sciences.

The clinical science component has 120 multiple-choice questions that examine a candidate’s clinical medicine skills, while the basic science section contains 120 questions that assess a candidate’s competency in the basic sciences of medicine. It is made up of multiple-choice questions. The TUS test is a significant intellectual challenge that necessitates a complete comprehension of the subject content, emphasizing the importance of excellent study and preparation strategies [14, 15]. The prospective role of AI and LLM becomes even more relevant in this context.

Despite potential limits and ethical concerns, the benefits of incorporating NLP and LLM into these fields are substantial. These technologies, for example, can boost student engagement, interaction, and customized learning, as well as help students score well on subject-specific assessments such as the TUS [16]. The digital revolution of healthcare, together with developments in AI technologies such as GPT-3.5 and GPT-4, point to a bright future for AI in medical education. However, the future must be handled properly, continually re-evaluated, and changed in response to changing social requirements, technical developments, and a growing awareness of AI’s capabilities and limits.

The goal of this study was to compare the performance of the GPT-4 and GPT-3.5 AI models in answering questions on the Turkish Medical Specialty Examination (TUS), with an emphasis on the differences between the clinical and fundamental science parts. This study is significant for both educators and AI developers because it gives a thorough grasp of the prospective uses of AI in medical education.

## 2. Materials and methods

### 2.1 Data Collection

This study analyzed data from Turkish Medical Specialties (TUS) examination records from the past three years, which included six TUS examinations and a total of 1440 medical questions. The Council on Higher Education (CoHE) publishes this data annually, and it contains question difficulty scores, selection scores, and key answers, assuring a balanced mix of questions from the science sections, basic and clinical studies.

### 2.2 Simulation Setup

Two AI models, GPT-4, and GPT-3.5, were charged with responding to collected TUS questions in simulations. They were instructed to select what they find to be the correct answer or to skip the question. Due to their limitations in interpreting visual data, the models were given the option to skip questions that included images.

### 2.3 Answer Scoring

The AI models’ responses were scored using answer keys provided by CoHE . The official TUS scoring system, which involves computing raw scores from correct and incorrect answers and then normalizing them into points, is used to provide a score after cross-checking each response with the primary response. specific T and K scores (for clinical and basic medical sciences, respectively).

### 2.4 Data Analysis

After evaluation, IBM SPSS 26.0 software was used to combine and analyze the data. Following a comparison of the overall results between GPT-4 and GPT-3.5, individual analyses for the basic science and clinical sections were conducted.

To get insight into how well each AI model performed, descriptive statistics (mean, mean, and standard deviation) of the scores were computed. Inferential statistics were applied to compare the performance of the two AI models. Independent sample tests or Mann-Whitney U tests were conducted, depending on how the data were distributed. In order to compare the performance of two models on the same set of questions, paired sample t-tests were also applied, where needed.

Analysis of variance (ANOVA) was performed to compare the model’s performance across courses. If ANOVA revealed significant differences, graduate tests were used to pinpoint individual courses where these discrepancies occurred.

Correlation analysis was also carried out to look into any potential connections between question difficulty, selectivity, and model performance. Regression analysis was used if more complex studies were required to forecast the model’s performance depending on these characteristics.

## 3. Results

### 3.1 GPT 3.5

The mean success rate across all domains was 40.17%, with 1 to 25 correct answers per test. The model gave 10.23 inaccurate answers and skipped 0.12 questions on average. The average number of correct answers for Clinical Medical Sciences was 63.17 (SD=3.19), with answers ranging from 58 to 67. Participants skipped 0.33 (SD=0.52) questions on average, with a range of 0 to 1. The average number of incorrect answers was 56.50 (SD=3.56), with a range of 52 to 62. The average net score was 49.04 (SD = 4.08), with a range of 42.50 to 54. The overall score varied from 47.43 to 53.05, with a mean of 50.91 (SD =1.89).

The average number of correct answers for Basic Medical Sciences was 63.00 (SD=7.75), with answers ranging from 50 to 73. With a range of 0 to 2, participants skipped an average of 1.17 (SD=0.75) questions. Incorrect answers ranged from 46 to 68, with an average of 56.00 (SD=7.29). With a range of 33 to 61.50, the average net score was 49.00 (SD=9.57). The overall score ranged from 47.08 to 55.29, with a mean of 52.05 (standard deviation=2.82).

### 3.2 GPT4

The overall success rate was 70.56%, with a range of 2 to 38 correct answers per test. The model gave 4.68 incorrect answers and skipped 0.12 questions on average. The average number of correct answers for Clinical Medical Sciences was 95.17 (SD=5.56), with answers ranging from 88 to 101. Participants skipped an average of 0.17 (SD=0.41) questions out of a possible total of 1. Incorrect answers ranged from 19 to 32, with an average of 24.67 (SD=5.75). The average net score was 89.00 (SD = 7.00), with a range of 80 to 96.25. The overall score varied from 62.90 to 74.49, with a mean of 70.40 (SD =4.26).

The average number of correct answers for Basic Medical Sciences was 92.00 (SD=7.51), with answers ranging from 78 to 99. With a range of 0 to 4, participants skipped an average of 1.17 (SD=1.60) questions. Incorrect answers ranged from 19 to 42, with an average of 26.83 (SD=8.13). With a range of 67.50 to 93.75, the average net score was 85.29 (SD=9.52). The overall score varied from 62.77 to 74.56, with a mean of 70.59 (SD =4.36).

### 3.3 Physicians

The participating physicians’ mean success rate was 38.14%, which is comparable to the GPT-3.5 model’s performance when compared to the AI models. Between 3.1 and 24.35 correct answers were given on average per test by participants. The average number of correct answers for Clinical Medical Sciences was 63.58 (SD=1.83), ranging from 60.44 to 65.77. The net score varied from 48.92 to 54.24, with a mean of 52.47 (SD =1.93).

The average number of correct answers for Basic Medical Sciences was 53.43 (SD=2.07), ranging from 51.63 to 57.34. The net score varied from 39.23 to 45.53, with an average of 41.10 (SD=2.32) (Table 1.).

**Table 1.**
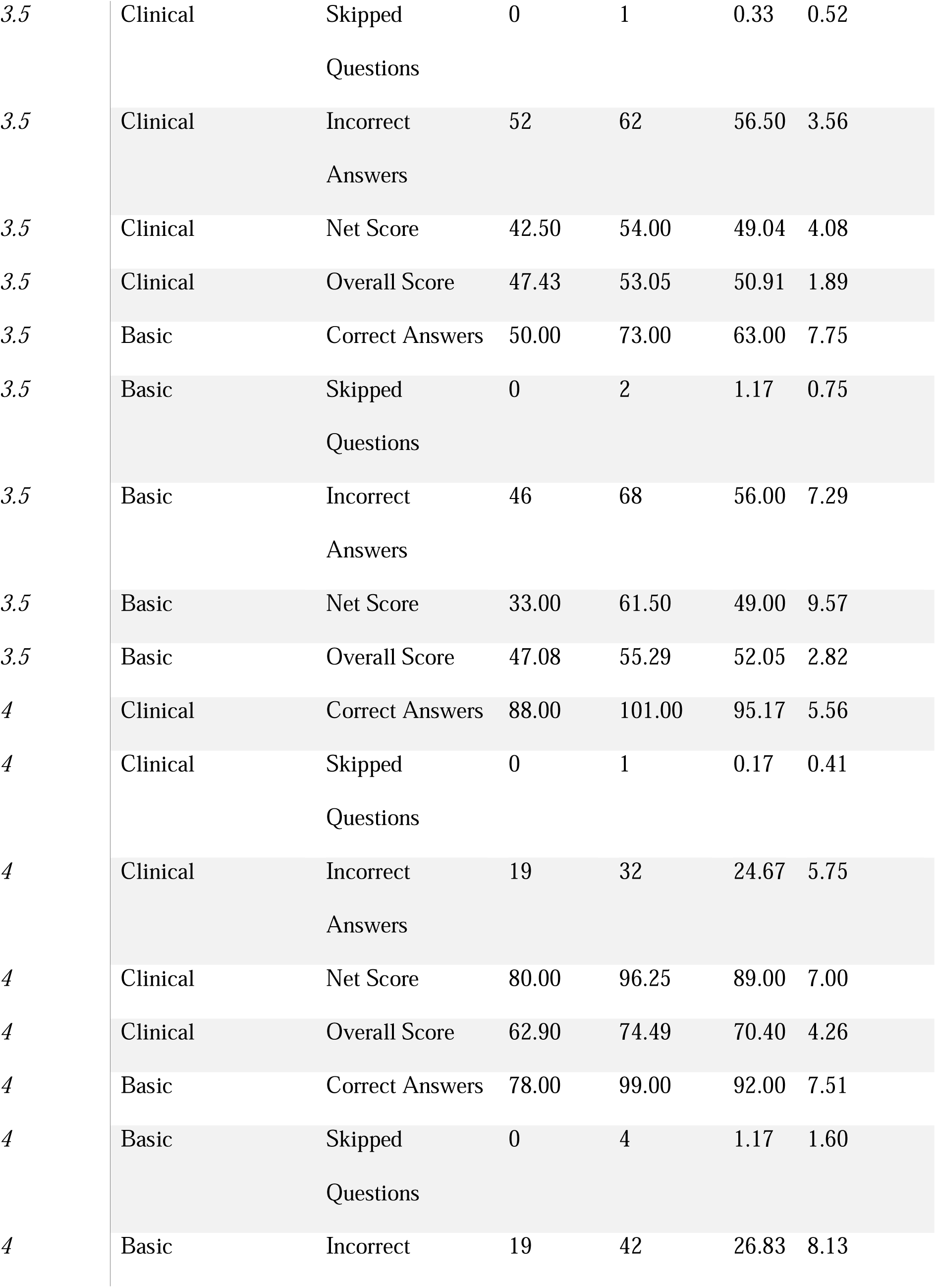
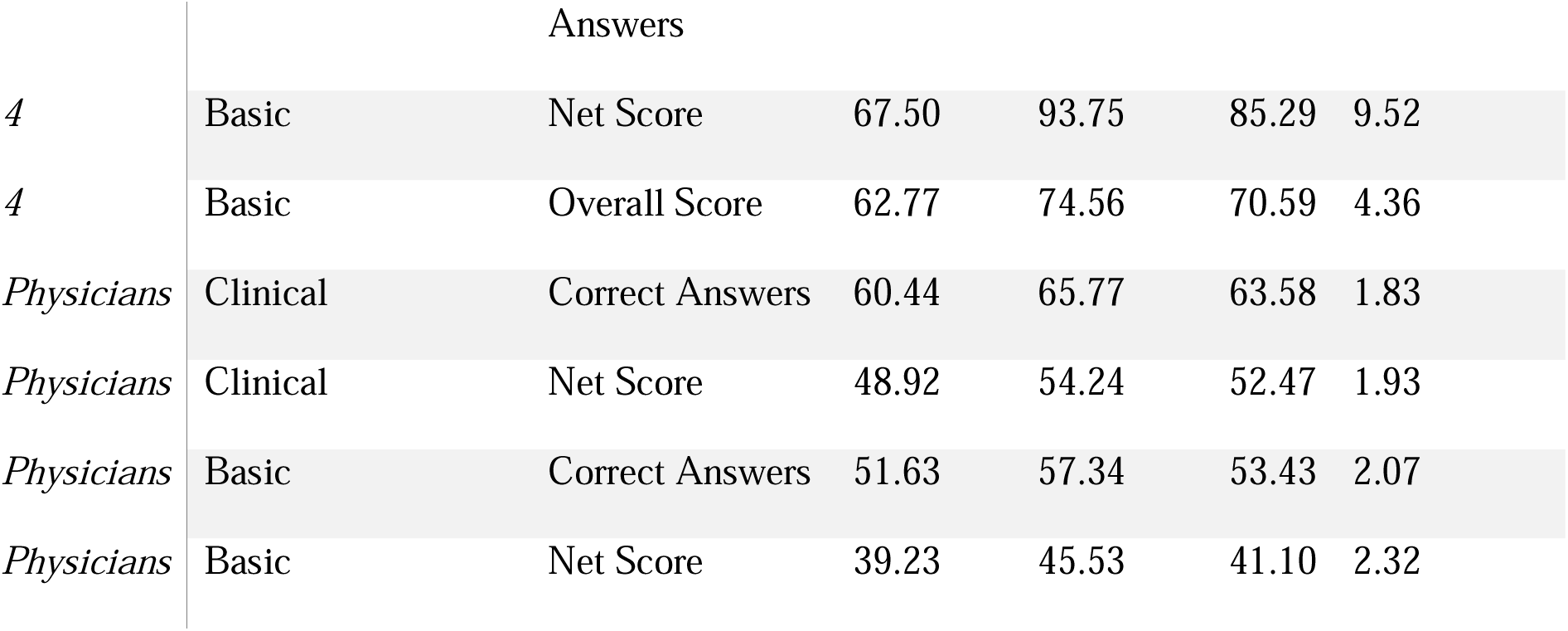
Descriptive Statistics of AI Models and Physicians Across Exams

The performance differences between OpenAI’s GPT-4 and GPT-3.5, as well as a comparison to the mean performance of 44,050 physicians, have been demonstrated by the results of the paired sample t-test.

First, there was a substantial difference in the correct answer counts of GPT-4 and GPT-3.5, with GPT-4 providing more correct responses (mean difference = 5.545, p<0.001). GPT-4 made much less inaccuracies as well (mean difference = 5.545, p<0.001). The number of questions skipped by the two AI models, however, did not differ significantly (p = 0.388).

Furthermore, the net scores (calculated by subtracting the incorrect response count divided by four from the correct answer count) for GPT-4 were substantially higher than for GPT-3.5 (mean difference = 6.93182, p<0.001).

When compared to the physician’s participants, GPT-4 had a larger number of accurate responses (mean difference = 6.37818, p<0.001) and a higher net score (mean difference = 7.33848, p<0.001).

Interestingly, GPT-3.5 outperformed the physicians in terms of correct answer count (mean difference = 0.8327, p = 0.033), although the difference in net scores was not statistically significant (p = 0.387).

Finally, the GPT-4, GPT-3.5, and medical doctor participants’ success rates (calculated score divided by number of questions) were examined. GPT-4 had a success rate of 70.56% on average, compared to GPT-3.5’s 40.17% and the participants’ 38.14%. The success rates differed statistically between GPT-4 and GPT-3.5, GPT-4 and the participants, and GPT-3.5 and the participants (Figure 1.) (Figure 2.).

**Figure 1.**
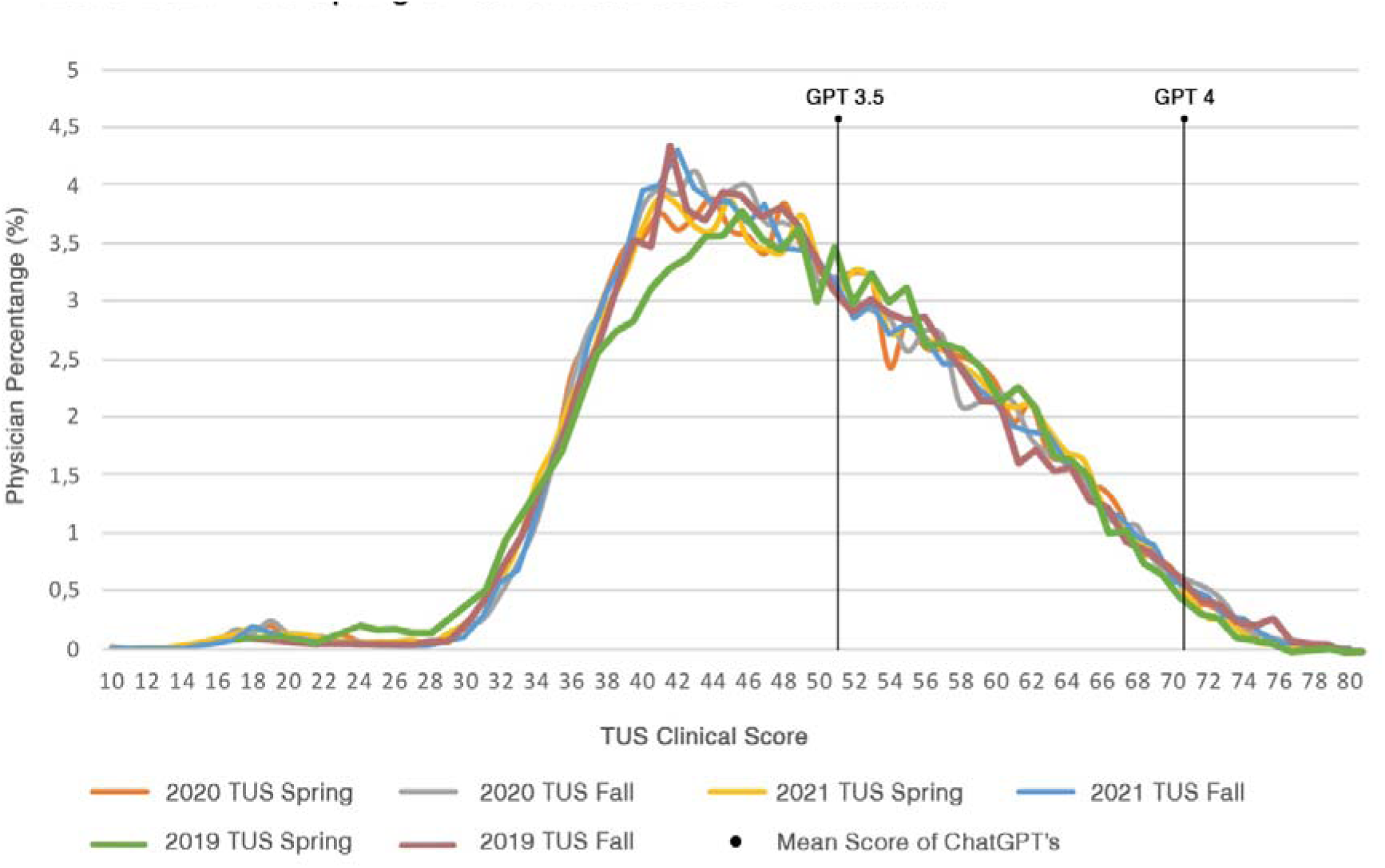
2019-2021 TUS Spring & Fall Basic Sciences Score Distribution

**Figure 2.**
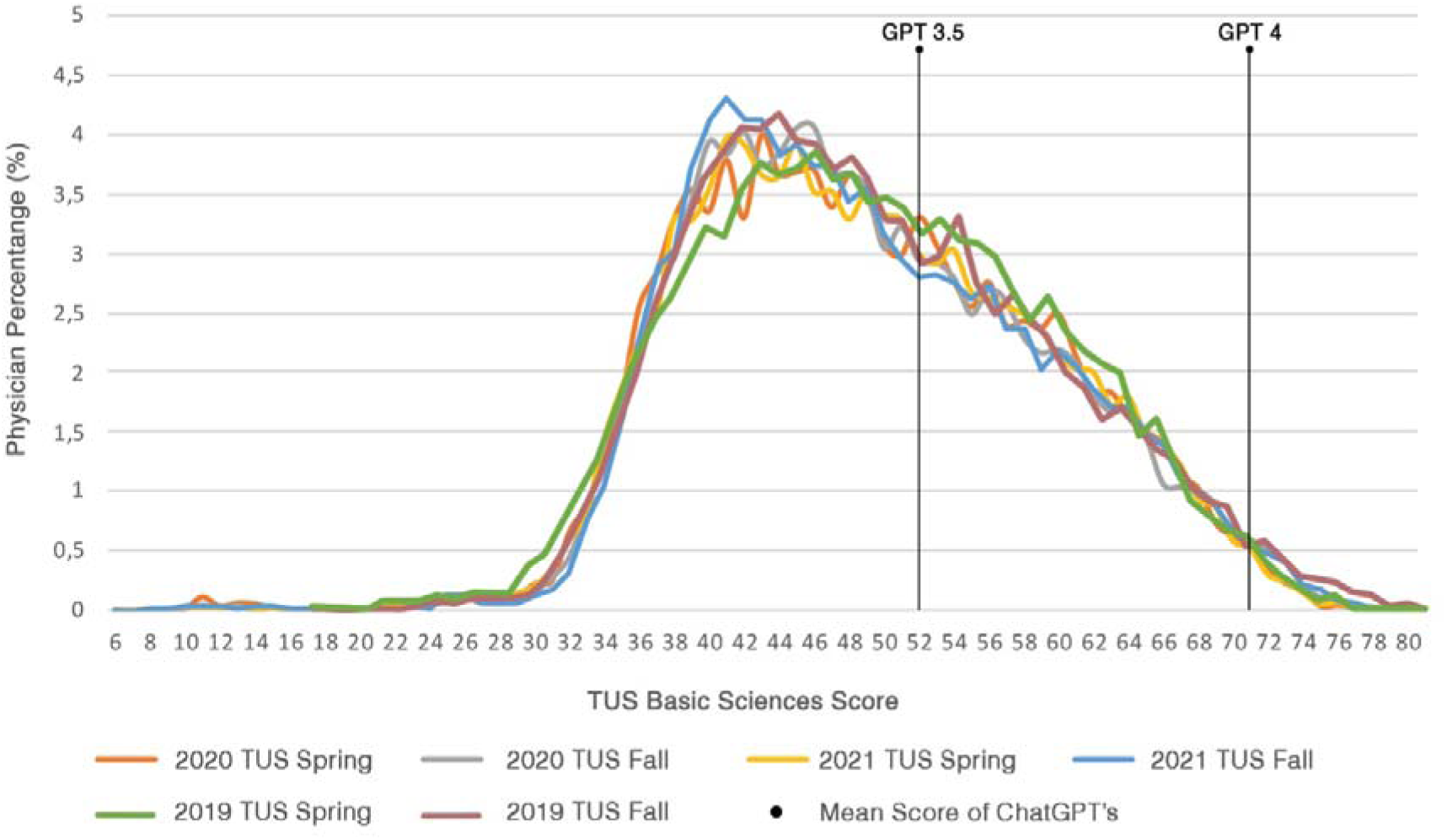
2019-2021 TUS Spring & Fall Clinical Score Distribution

A comparison analysis was performed in this study to compare the relative performance of AI models GPT-4 and GPT-3.5 against a cohort of 44,050 physicians across a variety of tests. Three unique performance ratio measures were used to facilitate this. The ’GPT3.5 Performance Ratio’ and ’GPT4 Performance Ratio’ were calculated by extracting the success rates of the physicians from the success rates of respective AI’s, allowing an evaluation of each AI’s relative effectiveness compared to the physicians. Whereas the ’GPT4 to GPT3.5 Performance Ratio’ was calculated by comparing the success rates of the two AIs, highlighting the variations in their effectiveness.

One-way ANOVA found significant differences between groups for both the ’GPT3.5 Performance Ratio’ (F(10, 55)=2.682, p=.010) and the ’GPT4 Performance Ratio’ (F(10, 55)=2.281, p=.026). The ’GPT4 to GPT3.5 Performance Ratio’, on the other hand, showed no significant variation (F(10, 55)=0.437, p=.922).

Further investigation utilizing Tukey HSD post-hoc comparisons revealed significant differences in the ’GPT3.5 Performance Ratio’ between the Anatomy and Pharmacology test groups, with a mean difference of -33.89 and a p-value of 0.037. The ’GPT4 Performance Ratio’ showed a comparable significant difference, with a mean difference of -33.74 and a p-value of 0.038.

Overall, the data indicate that the performance ratios of GPT-4, GPT-3.5, and Medical Doctors varied significantly depending on the field. The AI models outperformed the physicians in the pharmacology tests, while the physicians outperformed the AI models in the anatomy tests. Surprisingly, no significant difference in performance ratios was found between GPT-4 and GPT-3.5 across all disciplines.

Pearson’s correlation coefficient was used to analyze the correlations between the study’s variables. Most of these variables, including the difference in success rates between GPT4 and physicians, the GPT4 to GPT3.5 Performance Ratio, and the difference in success rates between GPT3.5 and physicians, did not demonstrate a significant link with the Average Discrimination Score. It only had a slightly negative association with the Average Difficulty Level (r = -.328, p < .05) (Table 2.).

**Table 2.**
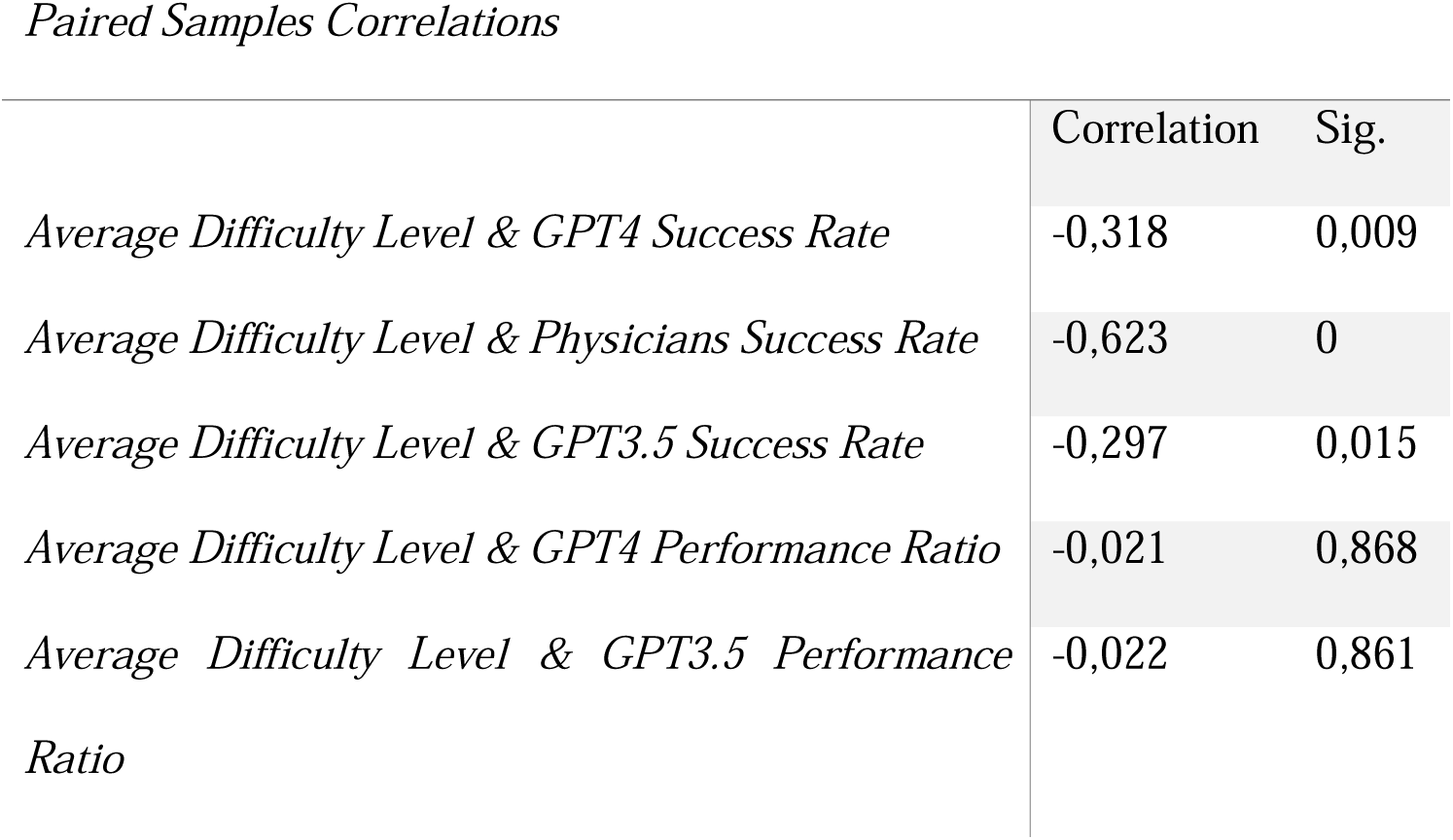
Paired Analysis of Average Difficult Level with Success Rates and Performance Ratios

Similarly, there was no significant link between the Average Difficulty Level and the difference in success rates between GPT4 and physicians, the GPT4 to GPT3.5 Performance Ratio, or the difference in success rates between GPT3.5 and physicians. However, there were substantial negative associations with both GPT4’s success rate (r = -.318, p < .01) and participant success rate (r = -.623, p < .01), indicating that as difficulty increases, both GPT4 and human participants’ success rates fall (Figure 3.).

**Figure 3.**
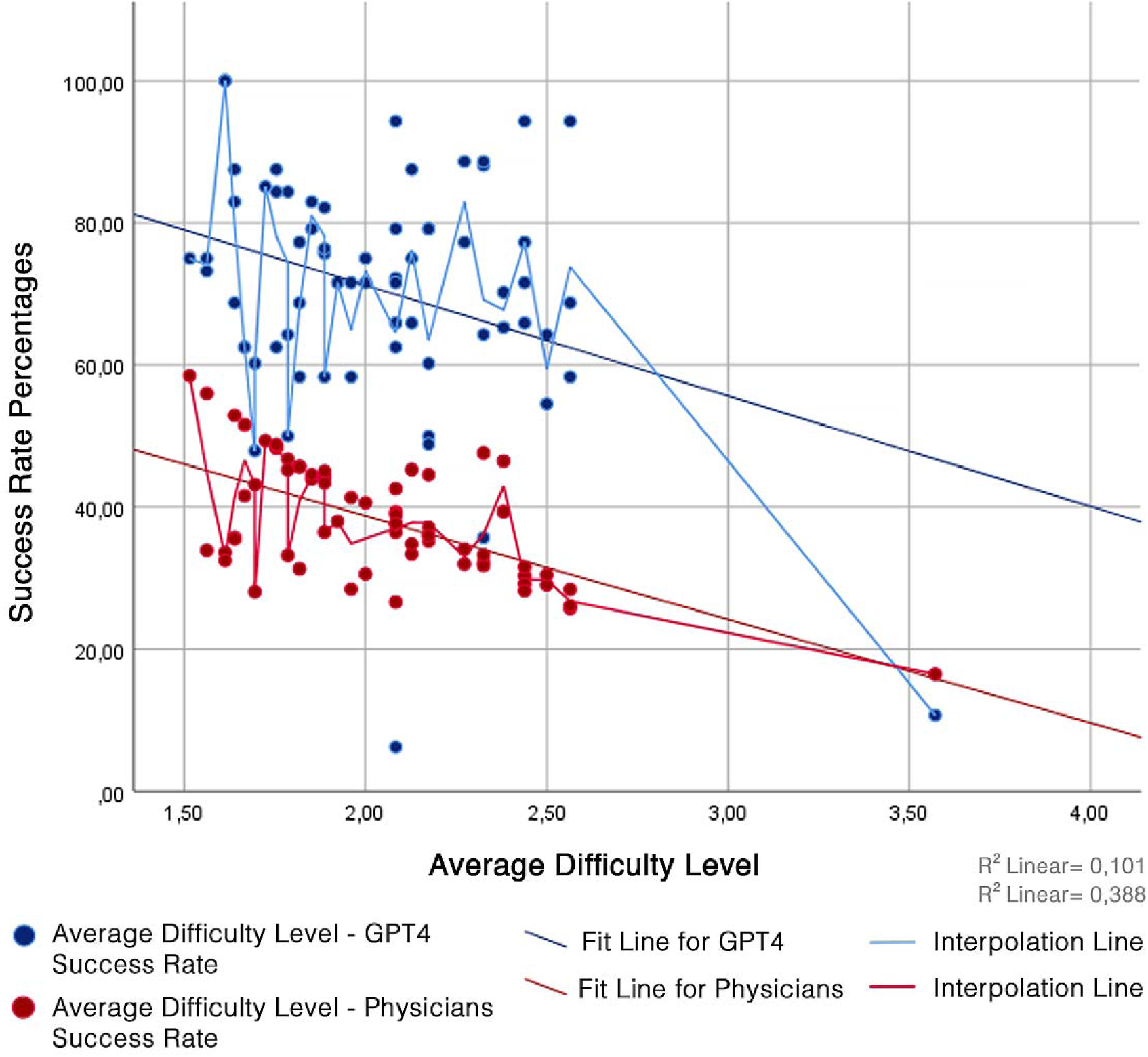
Average Difficulty Level & GPT4 Success Rate and Physicians Success Rate

## 4. Discussion

The findings represented a comparison of the performance of AI models GPT-4 and GPT-3.5 in a series of tests against a large sample of physicians. Correct answers, incorrect answers, skipped questions, net scores, and total success rates are all key performance measures. GPT-4 had a significantly greater success rate, averaging 70.56%, than GPT-3.5, which had a mean success rate of 40.17%. The participating physicians, on the other hand, had a 38.14% success rate, which was roughly the same as GPT-3.5. One-way ANOVA and Tukey HSD post-hoc analyses demonstrated a substantial difference in the performance ratio between the two AI models and the physicians, particularly in Anatomy and Pharmacology.

GPT-4 outperformed GPT-3.5 in several aspects, including producing more correct responses (5.545), fewer incorrect answers (6.545), and significantly higher net scores (6.633182). These distinctions indicate that GPT-4 has a deeper understanding of the subject matter and is more competent at applying medical information. The amount of questions skipped by both models, however, did not change considerably, implying that GPT-4’s innovations did not significantly increase its capacity to handle queries it did not instantly know how to answer. GPT-4 also outperformed GPT-3.5 in both Basic and Clinical Medical Sciences examinations, outperforming it by 30.39%. The study discovered no statistically significant differences in performance ratios across all disciplines, demonstrating that GPT-4’s excellence is consistent across all domains of medical science.

Further comparisons between the performance of the AI models and the participating physicians revealed intriguing findings. While GPT-3.5 scored comparable overall performance ratings to physicians, it significantly outperformed physicians in the number of right answers delivered, even if the difference in net scores was not statistically significant. This implies that, while GPT-3.5 may provide more correct answers than physicians, it may also provide more incorrect answers, indicating a lack of precision that may stem from the fact that, unlike human physicians, AI models lack the ability to reason beyond the information on which they were trained.

GPT-4 surpassed both GPT-3.5 and physicians, suggesting substantial advances in knowledge and precision. The substantial negative correlation between Average Difficulty Level and GPT-4 success rate and participant success rate implies that increased difficulty has an influence on both AI and human performance. This similarity emphasizes the idea that AI performance in the medical industry is comparable to human performance, reinforcing AI’s role as a beneficial tool in assisting physicians in difficult decision-making processes.

The study emphasizes the possibilities and limitations of AI models featuring GPT-4 and GPT-3.5 in medical education, particularly compared to human physicians. These models have limits in reasoning beyond supplied knowledge [17], which is critical in topics such as anatomy, where visualization and comprehension of spatial interactions between distinct body parts are required. The study also discovered a substantial negative association between test difficulty and AI performance, implying that as the complexity of the exam increases, it negatively impacts AI’s performance.

The GPT-4 model addressed medical questions with outstanding accuracy, making it a helpful tool for medical students. However, particular risks must be considered, such as the model’s tendency to deliver inaccurate replies and significant unpredictability in performance across different medical areas [18]. Furthermore, possible biases in AI performance testing must be carefully analyzed to guarantee that the model’s abilities are accurately interpreted.

Although AI models have the potential to improve learning, they are unable to replace human cognition or critical thinking. To overcome this, a blended learning strategy that allows human instructors to focus on developing critical thinking and problem-solving skills could be advantageous [19]. Furthermore, training for students to engage with and critically analyze AI models should be offered as part of the medical curriculum, promoting a symbiotic link between AI and medical teaching.

ChatGPT did not meet the pass criterion in scenarios such as the American Heart Association’s Basic Life Support and Advanced Cardiovascular Life Support exams [20]. However, its ability to generate relevant, detailed answers and its alignment with resuscitation guidelines over other AI systems highlighted its potential as a valuable self-learning tool. The results of these research are relevant to the United States Medical Licensing Examination (USMLE), where ChatGPT’s logical justification of answers, even wrong ones, hinted at its potential as an interactive instructional tool [21, 22].

Results from the use of AI techniques such as ChatGPT and GPT-4 to non-English language testing have been inconsistent. While ChatGPT stumbled on the Taiwanese Family Medicine Board Exam, with only 41.6% accuracy, and the Taiwanese Pharmacist Licensing Examination, scoring between 54.4% and 67.6%, GPT-4 showed potential by passing numerous years of the Japanese Medical Licensing Exams [23–25]. These findings emphasize the necessity for additional AI model optimization, especially when working with non-Latin characters. The trials also highlight the need for AI solutions that can accommodate different linguistic and cultural contexts and the need of using a variety of assessment techniques in healthcare settings.

In the standardized admissions exams used in the UK, ChatGPT showed promise in evaluating aptitude, critical thinking, problem-solving, reading comprehension, and aptitude, but it struggled with questions that required a lot of science and math [26]. This highlights the importance of AI’s continual progress and educated integration with traditional learning methodologies.

The potential of AI technologies like ChatGPT in transforming education is evident in various studies. Understanding their capabilities, limitations, and modifying instructional approaches is crucial for successful application. Future studies should explore personalized learning, curriculum revision, and educational content development. AI integration in education and examination contexts is a continuous trend, requiring strategic planning, pedagogical modifications, and continuous review to maximize benefits while minimizing risks. Despite challenges such as analyzing AI’s efficiency, managing technical complexities, and preventing misuse, AI integration is a growing trend that requires continued focus and innovation.

## 5. Limitations

Despite the promising findings, this study has some limitations. The inability of GPT-4 and GPT-3.5 models to address image-based questions is an important limitation. The models continued to respond relying upon the associated text and options despite being told to ignore such questions. Although the number of responses was small in size, they may have influenced the total performance results.

Inconsistencies in the models’ responses were also shown by asking the same questions again. When identical questions were introduced on many occasions, both models demonstrated a tendency to select different responses, showing a measure of unpredictability. To prevent this, each TUS exam was administered twice using both models, with questions producing varied results repeated a third time. This inconsistency, however, remains a severe restriction, particularly in high-stake scenarios such as medical examinations.

## 6. Conclusion

The study investigates the potential and limitations of large-scale language models such as GPT-4 and GPT-3.5 in medical education. GPT-4 surpasses physicians in solving Turkish Medical Specialization Exam (TUS) questions and especially in pharmacology questions, demonstrating the development of AI technology. However, the models encounter difficulties in anatomy tests, necessitating further development and human intervention. Although AI models have made outstanding advances in medical information comprehension and application, they still face problems in areas such as visualization, spatial understanding, and reasoning beyond available knowledge. GPT-4 demonstrated significant accuracy in answering complicated medical questions, indicating that the use of AI in medical education is promising.

However, risks such as incorrect replies and performance variability require continuous monitoring and expanding development. The study also underlines the importance of combining AI with human instruction so that AI may supplement human cognition in areas of high complexity or difficulty. AI solutions that are inclusive and economical are critical for future AI growth, ensuring that AI education tools are accessible and beneficial to various learners. Despite its limitations, AI’s involvement in medical education is a growing trend with interesting future learning prospects.

## Data Availability

The data that support the findings of this study are available from the corresponding author, upon reasonable request.

## Acknowledgment/disclaimers/conflict of interest

In the conduct of this research, GPT-4 and GPT-3.5 AI models played pivotal roles. They were used in managing TUS Exam questions and significantly facilitated the utilization of SPSS for data analysis. These models were helpful in identifying potential errors and suggesting methodological improvements.

It should be noted that all findings derived with the assistance of these AI models underwent rigorous cross-validation to ensure accuracy and reliability. While the AI models were utilized to rectify grammatical inaccuracies, they were not engaged in creating substantial content, thereby maintaining the integrity and authenticity of our research. This research greatly benefited from the inclusion of these AI models in the process.

## References

1- Kusunose K. Revolution of echocardiographic reporting: the new era of artificial intelligence and natural language processing. J Echocardiogr. 2023 Jun 13 https://doi.org/10.1007/s12574-023-00611-1

2- Cheng K, Guo Q, He Y, Lu Y, Gu S, Wu H. Exploring the Potential of GPT-4 in Biomedical Engineering: The Dawn of a New Era. Ann Biomed Eng. 2023 Apr 28; https://doi.org/10.1007/s10439-023-03221-1

3- Chan KS, Zary N. Applications and Challenges of Implementing Artificial Intelligence in Medical Education: Integrative Review. JMIR Med Educ. 2019 Jun 15;5(1):e13930. https://doi.org/10.2196/13930

4- J. Qadir, “Engineering Education in the Era of ChatGPT: Promise and Pitfalls of Generative AI for Education,” 2023 IEEE Global Engineering Education Conference (EDUCON), Kuwait, Kuwait, 2023, pp. 1–9, https://doi.org/10.1109/EDUCON54358.2023.10125121

5- Farrokhnia M, Banihashem SK, Noroozi O, Wals A. A SWOT analysis of ChatGPT: Implications for educational practice and research. Innovations in Education and Teaching International. 2023 Mar 27; https://doi.org/10.1080/14703297.2023.2195846

6- Gao CA, Howard FM, Markov NS, Dyer EC, Ramesh S, Luo Y, et al. Comparing scientific abstracts generated by ChatGPT to real abstracts with detectors and blinded human reviewers. NPJ Digit Med. 2023 Apr 26;6(1):75. https://doi.org/10.1038/s41746-023-00819-6

7- Sallam M. ChatGPT Utility in Healthcare Education, Research, and Practice: Systematic Review on the Promising Perspectives and Valid Concerns. Healthcare (Basel). 2023 Mar 19;11(6):887. https://doi.org/10.3390/healthcare11060887

8- Farrokhnia M, Banihashem SK, Noroozi O, Wals A. A SWOT analysis of ChatGPT: Implications for educational practice and research. Innovations in Education and Teaching International. 2023 Mar 27; https://doi.org/10.2196/48291

9- Lo CK. What Is the Impact of ChatGPT on Education? A Rapid Review of the Literature. Education Sciences. 2023 Apr 18;13(4):410. https://doi.org/10.3390/educsci13040410

10- Mallio CA, Sertorio AC, Bernetti C, Beomonte Zobel B. Large language models for structured reporting in radiology: performance of GPT-4, ChatGPT-3.5, Perplexity and Bing. Radiol med. 2023 May 29; https://doi.org/10.1007/s11547-023-01651-4

11- Lee P, Bubeck S, Petro J. Benefits, Limits, and Risks of GPT-4 as an AI Chatbot for Medicine. N Engl J Med. 2023 Mar 30;388(13):1233–9 https://doi.org/10.1056/nejmsr2214184

12- Alqahtani T, Badreldin HA, Alrashed M, Alshaya AI, Alghamdi SS, bin Saleh K, et al. The emergent role of artificial intelligence, natural learning processing, and large language models in higher education and research. Research in Social and Administrative Pharmacy. 2023 Jun; https://doi.org/10.1016/j.sapharm.2023.05.016

13- Öcek Z, Batı H, Sezer ED, Köroğlu ÖA, Yılmaz Ö, Yılmaz ND, et al. Research training program in a Turkish medical school: challenges, barriers and opportunities from the perspectives of the students and faculty members. BMC Med Educ. 2021 Dec;21(1) https://doi.org/10.1186/s12909-020-02454-1

14- Turan S, Üner S. Preparation for a Postgraduate Specialty Examination by Medical Students in Turkey: Processes and Sources of Anxiety. Teaching and Learning in Medicine. 2015 Jan 2;27(1):27–36. https://doi.org/10.1080/10401334.2014.979186

15- Toy S, Sumeyye Bakici R. Analysis of anatomy questions asked in Medical Specialization Exams in year 2000 and beyond. Med-Science. 2022;11(1):120. https://doi.org/10.5455/medscience.2021.10.325

16- Denny JC, Spickard A, Speltz PJ, Porier R, Rosenstiel DE, Powers JS. Using natural language processing to provide personalized learning opportunities from trainee clinical notes. Journal of Biomedical Informatics. 2015 Aug;56:292–9. https://doi.org/10.1016/j.jbi.2015.06.004

17- Sinha RK, Deb Roy A, Kumar N, Mondal H. Applicability of ChatGPT in Assisting to Solve Higher Order Problems in Pathology. Cureus. 2023 Feb 20; https://doi.org/10.7759/cureus.35237

18- Thirunavukarasu AJ, Hassan R, Mahmood S, Sanghera R, Barzangi K, El Mukashfi M, et al. Trialling a Large Language Model (ChatGPT) in General Practice With the Applied Knowledge Test: Observational Study Demonstrating Opportunities and Limitations in Primary Care. JMIR Med Educ. 2023 Apr 21;9:e46599 https://doi.org/10.2196/46599

19- Banihashem SK, Noroozi O, den Brok P, Biemans HJ, Kerman NT. Modeling teachers’ and students’ attitudes, emotions, and perceptions in blended education: Towards post-pandemic education. The International Journal of Management Education. 2023 Jul;21(2):100803. https://doi.org/10.1016/j.ijme.2023.100803

20- Fijačko N, Gosak L, Štiglic G, Picard CT, John Douma M. Can ChatGPT pass the life support exams without entering the American heart association course?. Resuscitation. 2023 Apr;185:109732. https://doi.org/10.1016/j.resuscitation.2023.109732

21- Kung TH, Cheatham M, Medenilla A, Sillos C, De Leon L, Elepaño C, et al. Performance of ChatGPT on USMLE: Potential for AI-assisted medical education using large language models. PLOS Digit Health. 2023 Feb 9;2(2):e0000198. https://doi.org/10.1371/journal.pdig.0000198

22- Gilson A, Safranek CW, Huang T, Socrates V, Chi L, Taylor RA, et al. How Does ChatGPT Perform on the United States Medical Licensing Examination? The Implications of Large Language Models for Medical Education and Knowledge Assessment. JMIR Med Educ. 2023 Feb 8;9:e45312. https://doi.org/10.2196/45312

23- Kasai J, Kasai Y, Sakaguchi K, Yamada Y, Radev D. Evaluating GPT-4 and ChatGPT on Japanese Medical Licensing Examinations. arXiv [csCL]. Published online 2023. https://doi.org/10.48550/arXiv.2303.18027

24- Weng T, Wang Y, Chang S, Chen T, Hwang S. ChatGPT failed Taiwan’s Family Medicine Board Exam. Journal of the Chinese Medical Association. 2023 Jun 9;Publish Ahead of Print https://doi.org/10.1097/jcma.0000000000000946

25- Wang Y, Shen H, Chen T. Performance of ChatGPT on the Pharmacist Licensing Examination in Taiwan. Journal of the Chinese Medical Association. 2023 May 25;Publish Ahead of Print https://doi.org/10.1097/jcma.0000000000000942

26- Giannos P, Delardas O. Performance of ChatGPT on UK Standardized Admission Tests: Insights From the BMAT, TMUA, LNAT, and TSA Examinations. JMIR Med Educ. 2023 Apr 26;9:e47737. https://doi.org/10.2196/47737

